# Systematics investigation of key drivers of lung adenocarcinoma: A focus on genes, pathways, and miRNAs

**DOI:** 10.1101/2024.11.09.24317046

**Authors:** Maryam Navaei, Fatemeh Karami, Aria Jahanimoghadam, Sara Zareei, Babak Khorsand

## Abstract

**Introduction:** Lung cancer remain a leading cause of cancer-related death, largely due to its asymptomatic progression in early stages and the development of drug resistance. Non-small cell lung cancer (NSCLC) accounts for 80% of all lung cancer cases, with lung adenocarcinoma (LUAD) being the most prevalent subtype. Despite advancements in treatment, the 5-year survival rate for LUAD remains low. Therefore, exploring gene networks may reveal novel therapeutic targets and pave the way for improved

**Method:** A comprehensive literature review was conducted across various databases containing multi- level genomic information. From this, a robust list of LUAD-related genes was curated. These genes were used to construct a weighted network based on KEGG pathway similarity. The network was subjected to clustering, hub gene detection, and gene ontology analysis. In parallel, a protein-protein interaction (PPI) network was constructed around these genes, which was further enriched with miRNA data to develop a gene-miRNA regulatory network.

**Results:** Following our analysis, 48 genes were identified as crucial to LUAD. Many of these genes, along with their corresponding miRNAs, were found to be either upregulated or downregulated in LUAD tissues. The hub genes and miRNAs identified are believed to play key roles in the initiation and progression of LUAD. Our network analysis highlighted PIK3CA, BRAF, EGFR, ERBB2, FGFR3, MTOR, and TP53, along with KRAS, MET, and FGFR2, as potential biomarkers. Additionally, miR-17-5p and miR-27a-3p, which are notably implicated in LUAD, emerged as novel biomarker candidates.

**Conclusion:** In conclusion, we employed a combination of bioinformatics techniques and database mining to derive a refined list of genes and miRNAs with high potential for further research in LUAD. We also identified core pathways that play a critical role in LUAD pathogenesis, providing a foundation for future studies aimed at developing more targeted therapeutic approaches.

## Introduction

Lung cancer is one of the Leading causes of death worldwide, often referred to as “silent killer” due to its asymptomatic nature in the early stages and the development of drug resistance in advanced stages [1, 2]. These factors make it difficult to detect and treat, contributing to the rising mortality rate. Lung cancer is classified into two main types, Small Cell Lung Cancer (SCLC), which accounts for 15% of cases, and Non-Small Cell Lung Cancer (NSCLC), making up the remaining 85% [2]. Among the subtypes of lung cancer, lung adenocarcinoma (LUAD) is the most common, affecting both smokers and non-smokers alike[3]. Key risk factors for lung cancer include exposure to radioactive gases, tobacco smoke (both first-hand and second-hand), and air pollution [2]. Within NSCLC category, LUAD is the most prevalent subtype, representing over 40% of these cases [4, 5].

Despite advancements in in treatment options, including immunotherapy and minimally invasive surgical techniques, the prognosis for LUAD remains poor. The 5-year overall survival rate stands at only about 17.4, with an average survival of just 15%. While therapies such as chemotherapy and chemoradiation have led to some improvements, the increase in survival rates over the past few decades has been minimal [4, 6–8].

Analyzing gene networks can provide crucial insights into the mechanisms underlying cancer, aiding in the development of more effective diagnostic tools and therapies [9, 10]. Currently, a combination of treatments -including surgical resection, radiotherapy, chemotherapy, genetic testing, immunotherapy, and targeted therapy- are used to treat lung cancer, showing varying degrees of effectiveness. However, significant challenges remain, such as tumor drug resistance, difficulties in early-stages diagnosis, variability in individual treatment responses, and genetic heterogeneity [11, 12]. Identifying key hub genes and designing therapies that target them could offer a promising approach to better manage tumors. Understanding the function of these genes could have a significant impact on overcoming drug resistance and inhibiting tumor growth and survival [13–15].

In the present study, we mined multiple databases to compile a comprehensive list of genes involved in various aspects of LUAD pathogenesis. The use of diverse databases strengthens the statistical power, resulting in a refined, robust set of genes that accurately represent LUAD. We have introduced a novel approach to constructing gene networks, where connections (edges) are based on shared biological processes or KEGG terms, rather than traditional physical interactions.

As illustrated in Fig. 1 this method enables the identification of functional gene modules, offering deeper insight into the underlying biological mechanisms. Additionally, it helps to pinpoint critical pathways that might be overlooked in networks built solely on physical interactions, potentially uncovering novel therapeutic targets and biomarkers.

**Fig. 1.**
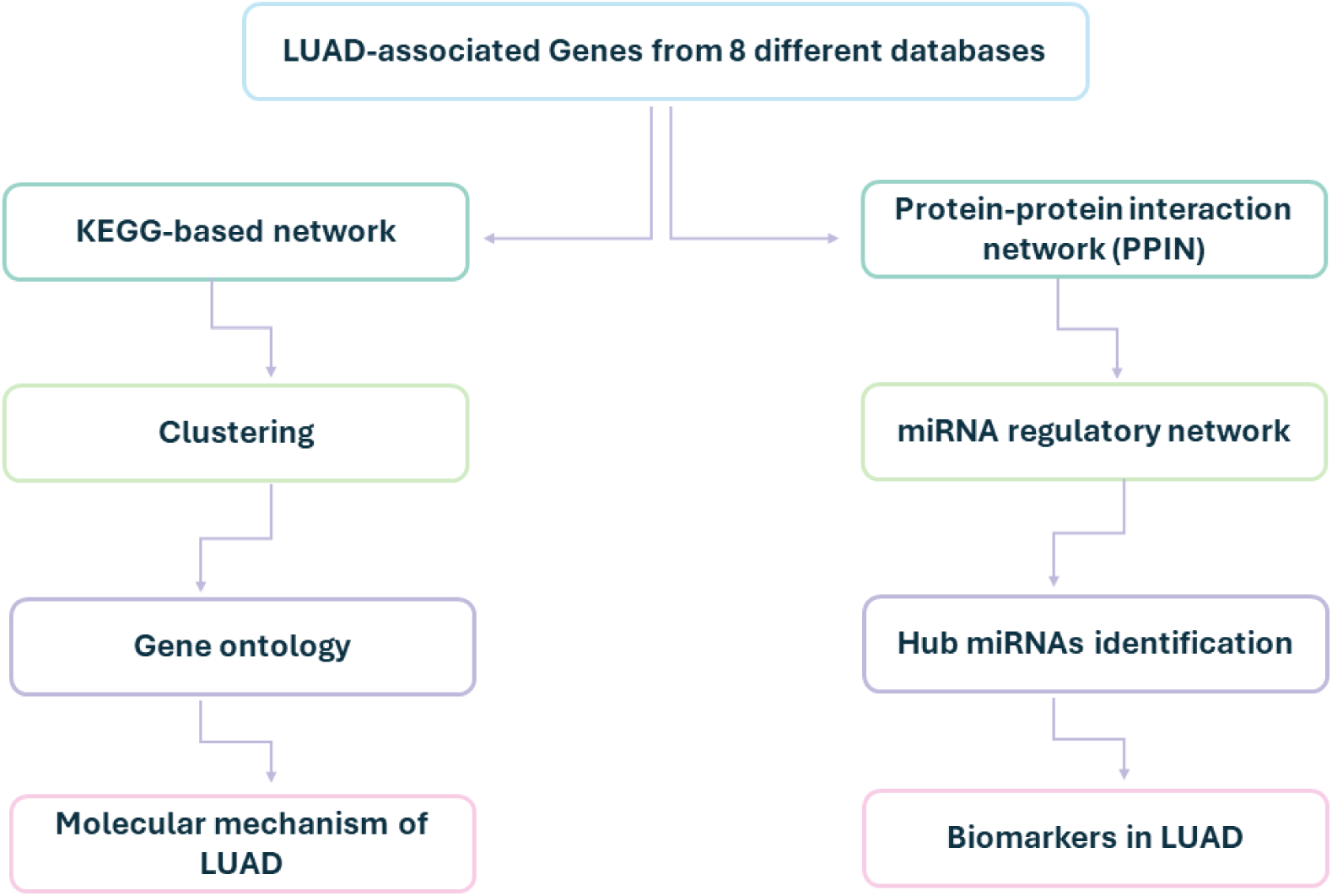
The flowchart illustrates methodology, starting from compilation of LUAD-associated genes to the identification of molecular mechanisms and potential biomarkers.

## Methodology

### Compiling a Robust Gene List for LUAD

The collected genes were sourced from eight databases: dbGaP [16], TCGA [17], Disgenet [18], OpenTarget [19], UniProt [20], CancerHotspot [21], and ClViC [22]. LUAD-related genes were extracted from each database to form a ranked list. Genes appearing in at least 3 databases, or top- ranked genes from a single dataset, were included even if they were not present in the minimum 3 datasets. Specific selection criteria for each database were as follows:

- dbGaP: Genes occurring more than seven times.
- TCGA: scores were normalized between 0 and 1, and genes with scores above 0.3 were chosen.
- Disgenet: genes linked to more than 1500 diseases or with a gene-disease association score (GDA) greater than 0.3 were filtered, with genes scoring above 0.4 selected.
- OpenTargets: 5 factors were considered: ClinVar, global score, cancer biomarker, ClinVar somatic, and Cancer Gene Census, with a score threshold of 0.05. The top 20% of genes were selectd
- UniProt, CancerHotspot, and CIViC: As no specific score were available, full gene lists were used to calculate the frequency.

This approach ensures a comprehensive gene list, capturing gens with different biological relevance across the databases.

### KEGG Term Assignment for Genes Using Enrichr

Enrichr [23] was used to perform enrichment analysis, determining biological significance for our final list of 48 genes. The gene list was used as an input into the Enrichr web server to obtain the relevant KEGG terms. The resulting KEGG terms were split for each gene, forming a data frame linking genes to their corresponding KEEG pathways.

### Calculating Edge Weights

An edge was created between gene pairs if they shared a KEGG term. The edge weights represent the strength of gene pair’s relationship, based on shared biological process, and are referred to as “KEGG-based network” hereafter.

### Network Construction Based on Common KEGG Terms

Using the “igraph” package in R version 4.4.1, we built networks where edges represented shared KEEG terms. This novel network construction method focuses on shared functional roles, offering deeper understanding of gene interactions within the LUAD context.

### Clustering Using MCODE Plugin in Cytoscape

Once the network was built, it was imported into Cytoscape version 3.10., a bioinformatics tool for network visualization and analysis. The MCODE plugin was used to identify highly interconnected modules (clusters) within the network, using default parameters (node score cutoff: 0.2, K-Core: 2).

### Gene Ontology (GO) Analysis

Gene Ontology and pathway enrichment analysis were performed to investigate the main functions and pathways in each cluster. Using gProfiler tool within Cytoscape, we indentified relevant GO terms, providing insight into how each cluster contributes to LUAD pathogenesis.

### Network Construction Based on Physical Interactions

To complement the KEEG-based network, we built a protein-protein interaction network (PPIN) using the STRING database (https://string-db.org/). Interactions were filtered using a minimum interaction score of 0.7, ensuring high-confidence interactions between genes.

### miRNA-Gene Regulatory Network Construction

A regulatory network around the PPIN was constructed using CyTargetLinker plugin in Cytoscape [24]. miRTarBase [25] was used as the source of miRNA-gene interactions. We identified the top 10 hub miRNA by ranking them based on their connectivity within the network.

## Results

A total of 48 LUAD-related genes were obtained from multiple databases, details of which can be found in Table S.1, including the frequency of each gene across the eight databases. Using KEGG terms derived from Enrichr and our defined methodology for establishing edges between genes, 29 of these genes shared at least one KEGG term. These 29 genes were used to construct the KEGG-based gene network.

To identify hub genes, 12 different centrality algorithms were applied within Cytoscape: CC, DMNC, MNC, Degree, EPC, Bottleneck, Eccentricity, Closeness, Radiality, Betweenness, Stress and Clustering Coefficient. For each algorithm, 10 hub genes were identified (Fig. 2, Table. 1). The most frequently recurring genes across these algorithms were PIK3CA, BRAF, EFGR, ERBB2, FGFR3, KRAS, MET, MTOR, TP53 and FGFR2, with PIK3CA being identified as a hub gene in 10 out of 12 algorithms. The network analysis also resulted in two distinct clusters: Cluster 1 with 22 nodes and 193 edges, and Cluster 2 with 3 nodes and 3 edges (Fig 3).

**Fig. 2.**
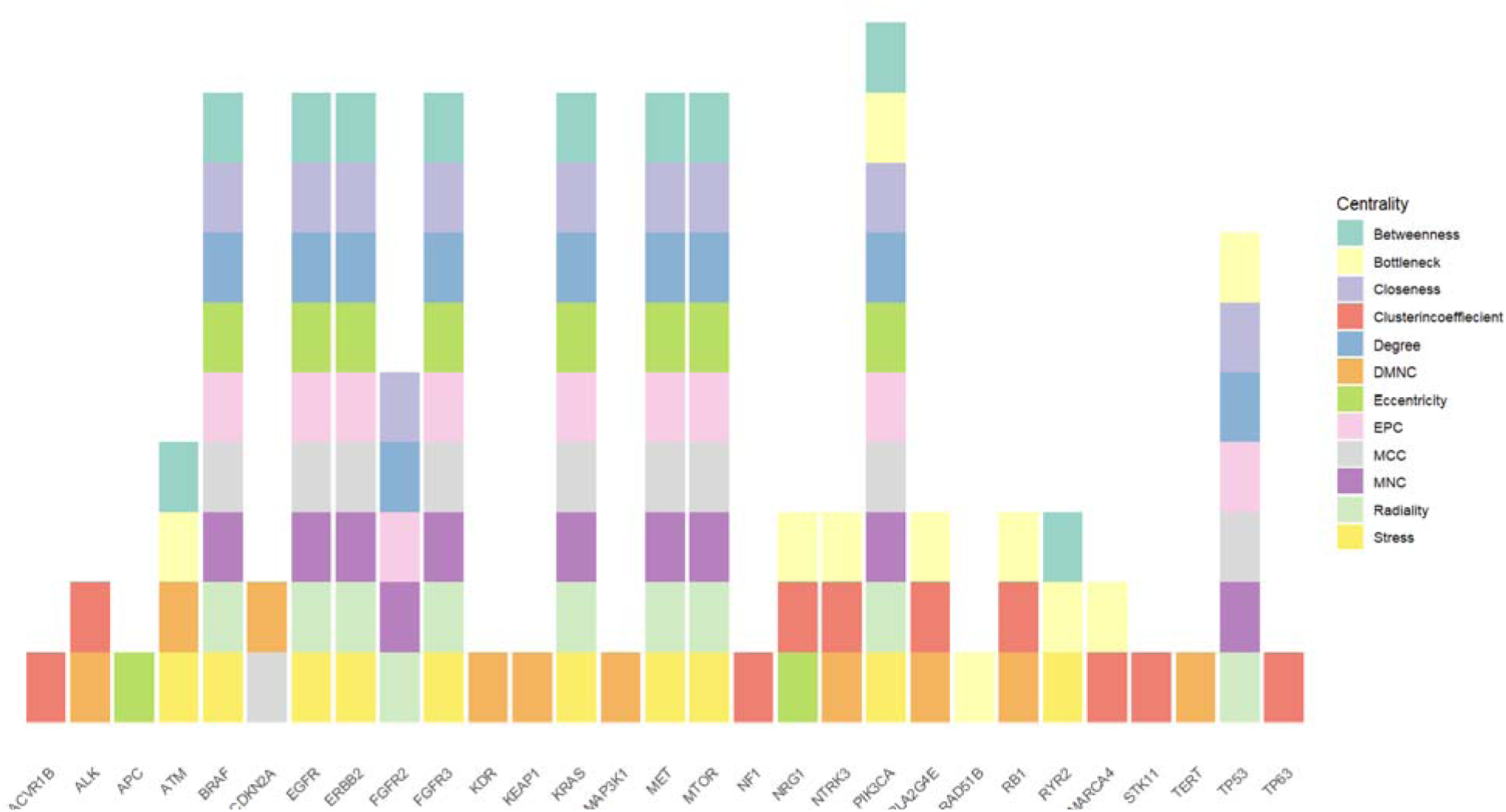
Frequency of hub genes identified across 12 different centrality algorithms

**Fig. 3.**
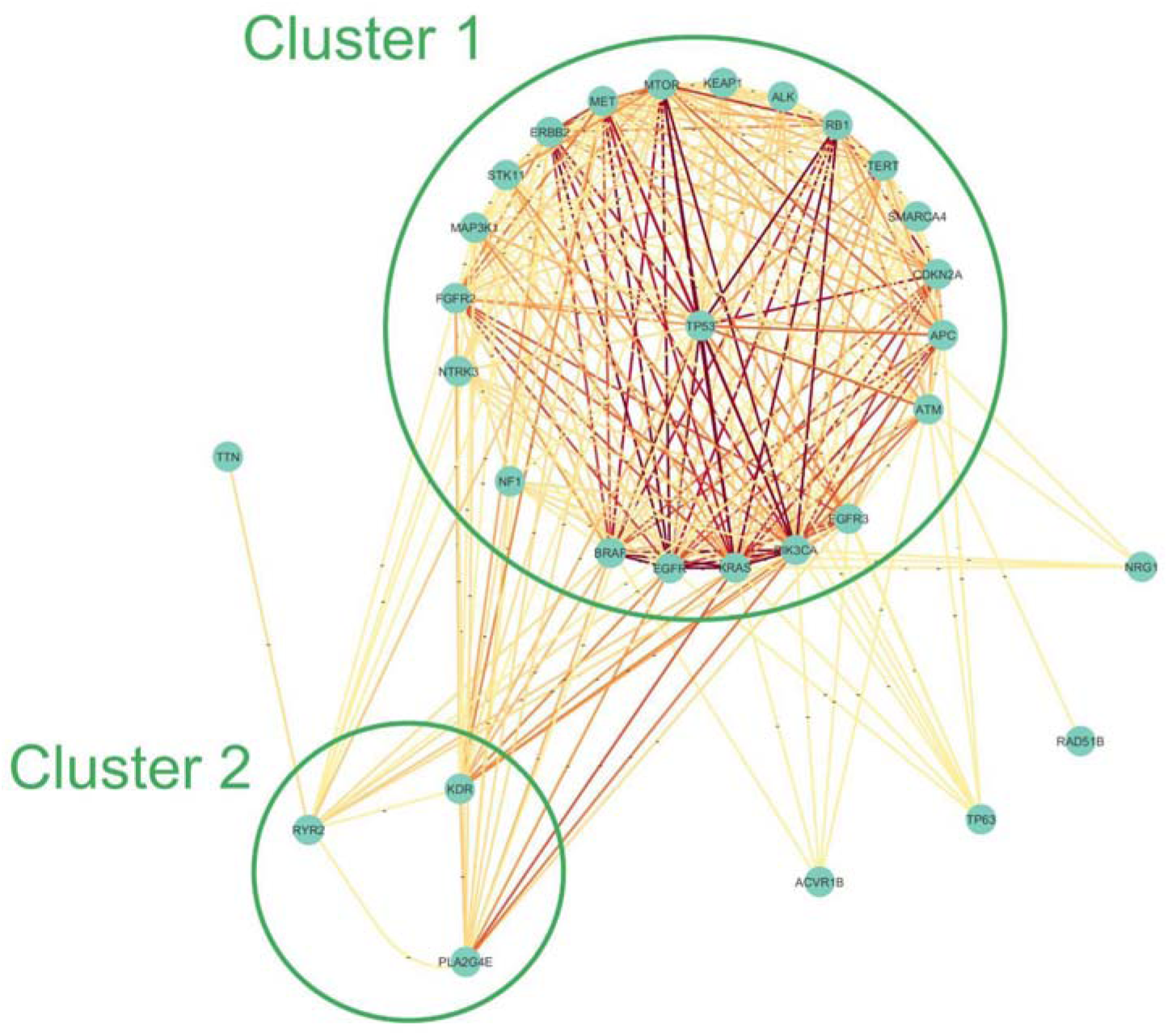
Two clusters identified from the KEGG-based network. Redder edges indicate stronger pathway dependencies between gene pairs.

**Table 1.** Frequency of hub genes among 12 different hub detection algorithms.

| Gene | Frequency | Gene | Frequency | Gene | Frequency |
| --- | --- | --- | --- | --- | --- |
| PIK3CA | 10 | NRG1 | 3 | KEAP1 | 1 |
| BRAF | 9 | NTRK3 | 3 | MAP3K1 | 1 |
| EGFR | 9 | PLA2G4E | 3 | NF1 | 1 |
| ERBB2 | 9 | RB1 | 3 | RAD51B | 1 |
| FGFR3 | 9 | RYR2 | 3 | STK11 | 1 |
| KRAS | 9 | ALK | 2 | TERT | 1 |
| MET | 9 | CDKN2A | 2 | TP63 | 1 |
| MTOR | 9 | SMARCA4 | 2 |  |  |
| TP53 | 7 | ACVR1B | 1 |  |  |
| FGFR2 | 5 | APC | 1 |  |  |
| ATM | 4 | KDR | 1 |  |  |

### Gene Ontology (GO) Analysis of KEGG-Based Clusters

The two clusters were subjected to GO analysis to further investigate the roles of specific genes in LUAD pathogenesis. Cluster 1 genes were significantly enriched in multiple biological process (BP), molecular function (MF), cellular component (CC) terms, as well as Reactome pathways (table. S.2, Fig. 4). Cluster 2 (Fig. 5) was associated with a single GO term, BP.

**Fig. 4.**
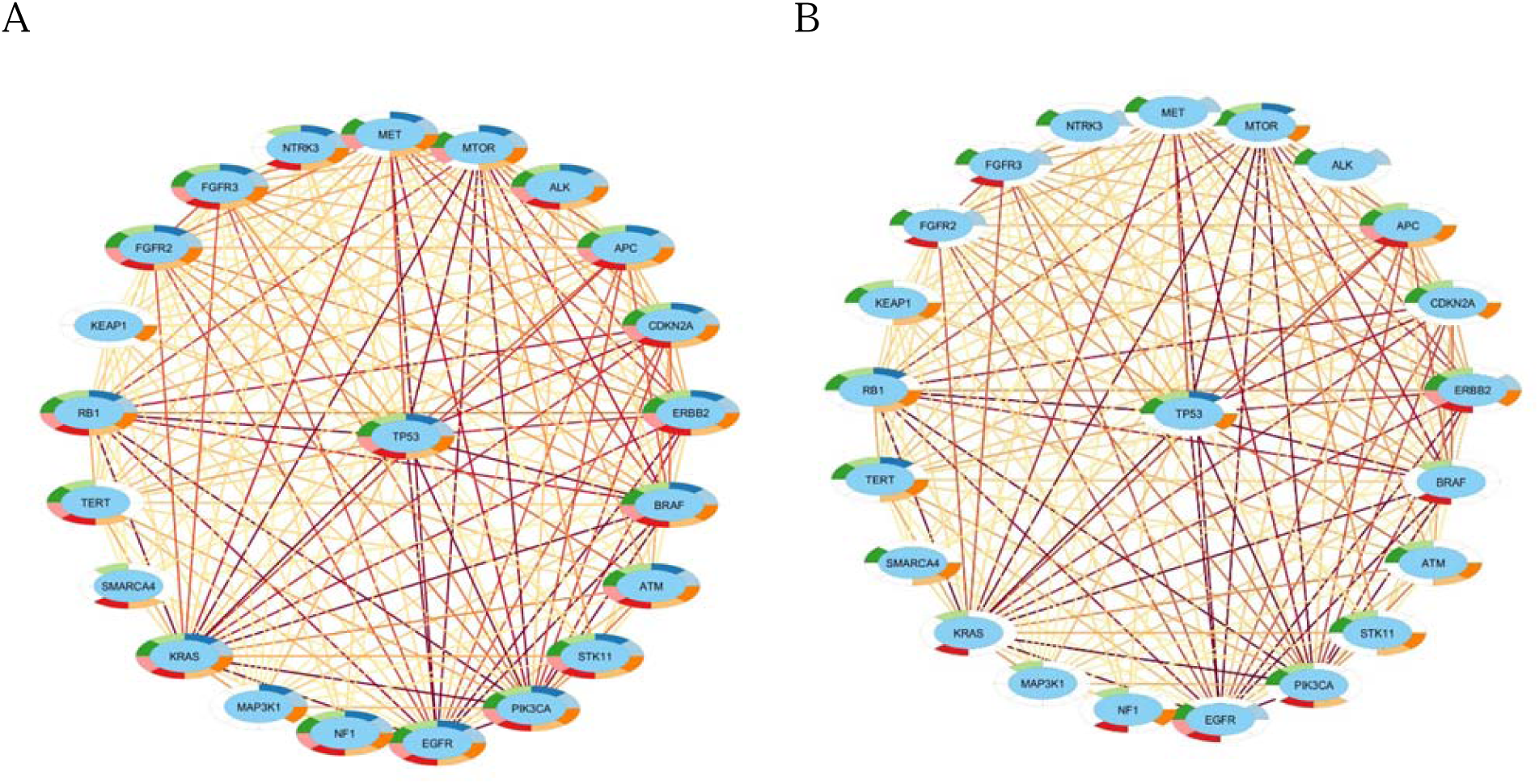
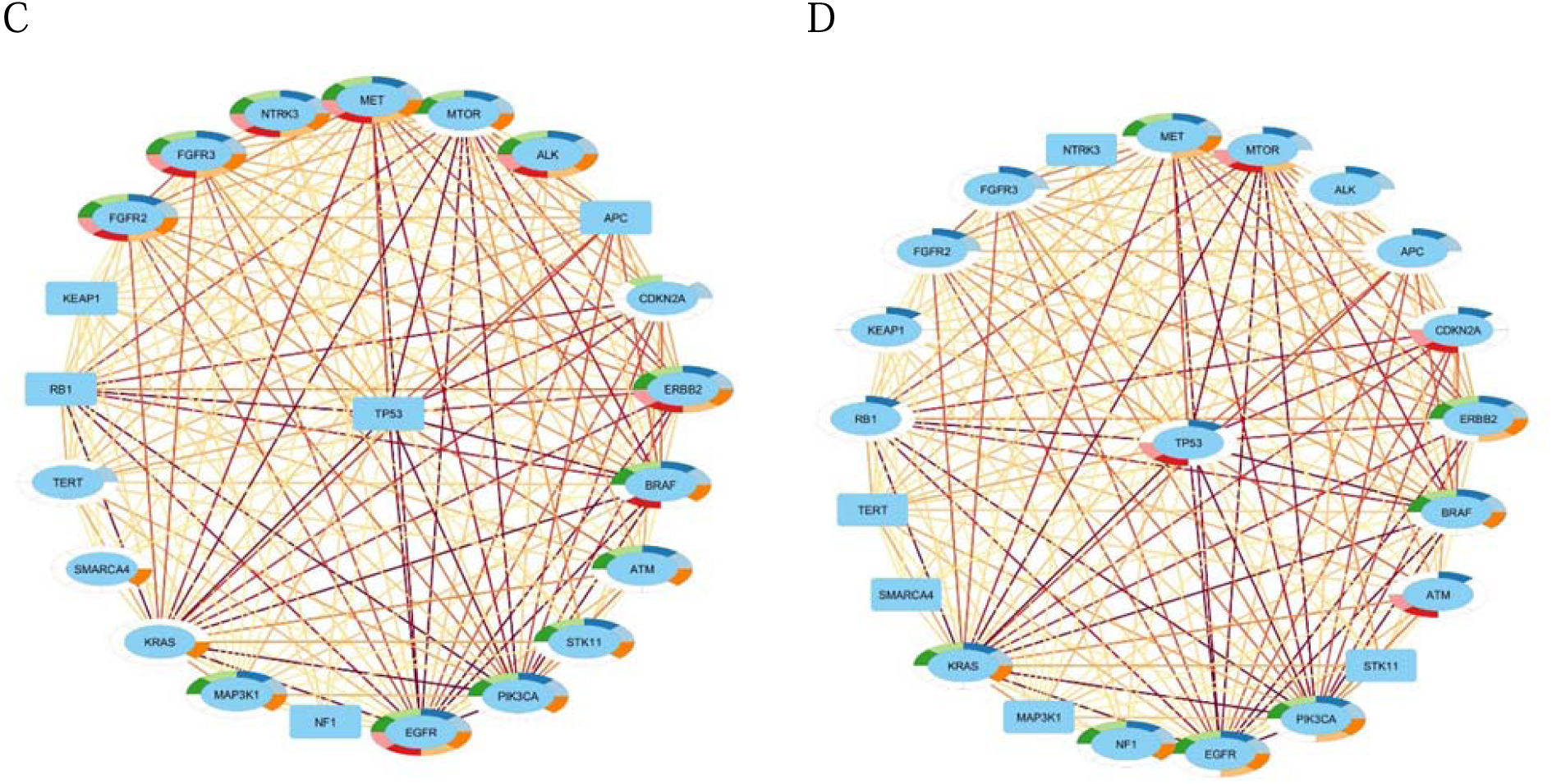
GO analysis results for genes in Cluster 1. (A) biological pathways, (B) cellular components, (C) molecular functions, and (D) Reactome pathways.

**Fig. 5.**
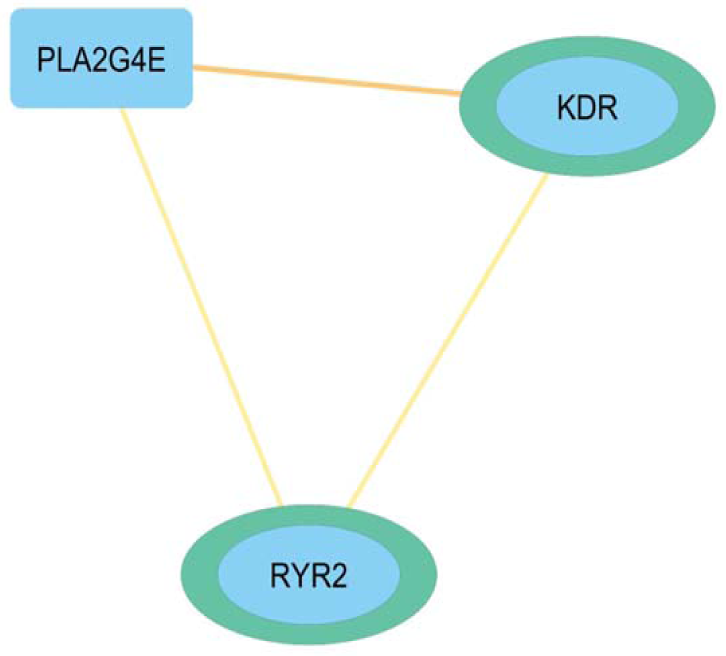
GO Analysis results for Cluster 2 genes.

For Cluster 1, the most significantly enriched biological process terms included: protein phosphorylation (GO:0006468; P=3.93E-17), phosphorylation (GO:0016310; P=8.04E-16), regulation of cell population proliferation (GO:0042127; P=1.49E-13), programmed cell death (GO:0012501; P=2.75E-12), cell population proliferation (GO:0008283; P=3.34E-12), regulation of signal transduction (GO:0009966; P=3.54E-12), protein modification process (GO:0036211; P=4.07E-12), and cell death (GO:0008219; P=2.85E-12).

The most notably enriched molecular function terms were: transferase activity, transferring phosphorus-containing groups (GO:0016772; P=5.10E-13), protein kinase activity (GO:0004672; P=1.15E-12), kinase activity (GO:0016301; P=1.29E-12), phosphotransferase activity, alcohol group as acceptor (GO:0016773; P=1.07E-11), transmembrane receptor protein tyrosine kinase activity (GO:0004714; P=1.09E-10), protein tyrosine kinase activity (GO:0004713; P=6.54E-10), transmembrane receptor protein kinase activity (GO:0019199; P=7.39E-10), and purine ribonucleoside triphosphate binding (GO:0035639; P=7.71E-9).

For cellular components, significant terms were: receptor complex (GO:0043235; P=9.09E-5), PML body (GO:0016605; P=6.44E-4), cytosol (GO:0005829; P=4.06E-3), protein-containing complex (GO:0032991; P=8.62E-3), ruffle membrane (GO:0032587; P=2.36E-2), cell junction (GO:0030054; P=2.83E-2), catalytic complex (GO:1902494; P=3.56E-2), and nucleoplasm (GO:0005654; P=3.98E-2).

Top Reactome pathways included: diseases of signal transduction by growth factor receptors and second messengers (REAC:R-HSA-5663202; P=1.33E-9), disease (REAC:R-HSA-1643685; P=1.41E-7), RAF/MAP kinase cascade (REAC:R-HSA-5673001; P=1.90E-4), MAPK1/MAPK3 signaling (REAC:R-HSA-5684996; P=2.15E-4), regulation of TP53 degradation (REAC:R-HSA-6804757; P=2.33E-4), regulation of TP53 expression and degradation (REAC:R-HSA- 6806003; P=2.61E-4), PI3K/AKT signaling in cancer (REAC:R-HSA-2219528; P=3.01E-4), and MAPK family signaling cascades (REAC:R-HSA-5683057; P=5.35E-4) (Fig. 6).

**Fig. 6.**
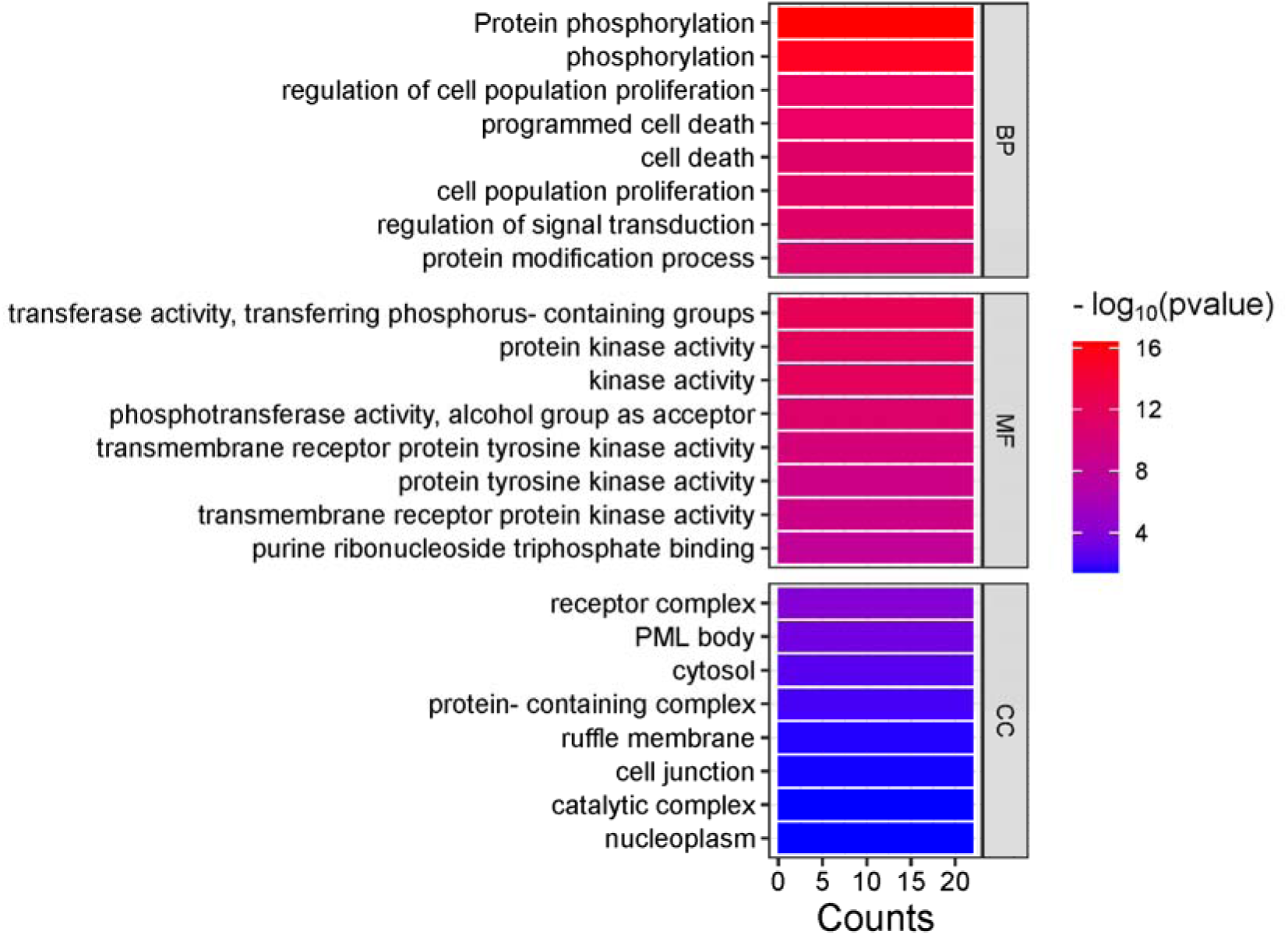
The results of GO analysis on cluster 1, illustrating the significance of various biological processes (BP), molecular functions (MF), and cellular components (CC). The heatmap uses colors ranging from blue to dark red to indicate different levels of significance (-log10(p value)), while the bar graph shows the counts for each category which is 22.

The most significantly enriched GO term for Cluster 2 was calcium-mediated signaling using intracellular calcium source (GO:0035584; P=1.58E-8).

### Protein-Protein Interaction Network (PPIN) and miRNA-Gene Regulatory Network

The PPIN constructed around our gene list consisted of 46 nodes and 109 edges, with two non-coding genes (LINC01511 and PHF5GP) excluded. Base on the PPIN, miRNAs were identified, with the top 10 being miR-335-5p, miR-16-5p, miR-125b-5p, miR-21-5p, miR-17-5p, miR-27a- 3p, miR-124-3p, miR-26b-5p, miR-155-5p, and miR-125a-5p (Table. 2)

**Table 2.** Literature review of top 10 miRNAs.

| miRNA | Degree | Regulation | Role as | Function in LUAD | Known as biomarker in LUAD |
| --- | --- | --- | --- | --- | --- |
| miR-335-5p | 10 | upregulated | oncomiR, tumor suppressor depending the target | cell proliferation, metastasis [26, 27] | Yes [28] |
| miR-16-5p | 8 | downregulation, upregulated | tumor suppressor | activating oncogenic pathways, diactivating tumor growth inhibiting pathways [29, 30] | Yes [30] |
| miR-125b-5p | 8 | downregulated | tumor suppressor | cell proliferation, metastasis [31] | Yes [32] |
| miR-21-5p | 7 | upregulated | oncomiR | cell proliferation, metastasis [33, 34] | Yes [33] |
| miR-17-5p | 7 | upregulated | oncomiR, tumor suppressor (in some context) | cell proliferation, metastasis [35, 36] | Not mentioned precisely |
| miR-27a-3p | 7 | upregulated | oncomiR | tumor development, metastasis, EMT, resistance to chemotherapy [37] | Not mentioned precisely |
| miR-124-3p | 6 | downregulated | tumor suppressor | metastasis, cellular metabolism, autophagy, and cell cycle regulation [38] | Yes [39] |
| miR-26b-5p | 6 | downregulated | tumor suppressor | cell proliferation, apoptosis [40] | Yes [41] |
| miR-155-5p | 6 | upregulated | oncomiR | cancer cell survival, progression, EMT [42] | Yes [43] |
| miR-125a-5p | 6 | downregulated | tumor suppressor | proliferation, invasion, apoptosis [44, 45] | Yes [32] |

## Discussion

The diagnosis and treatment of lung adenocarcinoma (LUAD) continue to face significant challenges, as evidenced by the increasing incidence of cases each year. Effective treatment strategies require a deeper understanding of LUAD pathogenesis and identification of reliable biomarkers. This study utilized multiple databases to identify key genes implicated in LUAD and shed light on the molecular mechanisms driving the disease.

Our analysis revealed PIK3CA as the most connected gene in the network, supported by its established correlation with poor survival in NSCLC patients. PIK3CA mutations occurs in 1.5% to 7.7% of LUAD cases and are involved in the PI3K/AKT1/MTOR pathway, often co-occurring with mutations in EGFR, KRAS, and BRAF [46–48].

Similarly, BRAF is another significant genes involved in kinase fusions and MAPK signaling, contributing to oncogenesis [49]. BRAF mutations, present in 3-5% of NSCLC cases, frequently co-occur with other mutation, TP53, KRAS, EGFR, NF1, STK11 and MET [50], and it has been identified as a biomarker for LUAD [46, 51].

EGFR, a receptor tyrosine kinase which is also known as ERBB1 and HER1, plays a role in growth factor signaling by promoting cell proliferation and inhibiting apoptosis [52]. EGFR is mutated in a substantial proportion of LUAD cases, with frequencies varying between populations. For instance, the mutation rate can reach up to 60%, in Asian populations. For example, a study conducted in China identified an EGFR mutation rate of 49.3% among adenocarcinoma patients, while another study reported a rate of 51.4% in a larger group of patients with advanced lung cancer [53, 54]. However, this value is generally lower (10-20%) in European and North American groups. EGFR mutations often co-exist with other genetic alterations like TP53, CDKN2A/B, BRCA2 and PTEN [55].

ERBB2 (HER2), another receptor tyrosine kinase, functions similarly to EGFR and is involved in PI3K/AKT, MAPK/ERK, JAK/STAT, and SRC/YAP pathways [56]. ERBB2 mutations, present in 1.5% to 2.6% of LUAD cases [57], often co-occur with mutations in TP53 [58], KRAS [59], PIK3CA [60] and FOXA1 [58], influencing tumor progression and treatment response.

In addition, FGFR3 mutations, although less frequent (around 1.3% of LUAD cases) [61], play a crucial role in regulating tumor growth and survival [31] by interacting with ley pathways, including JAK/STAT, MAPK and PI3K/AKT. Other genetic alterations like KRAS, PI3K, EGFR are often associated with FGFR3 mutations [61].

KRAS mutations, present in approximately 30% of LUAD cases, are particularly noteworthy due to their impact on various cellular pathways [62]. KRAS mutations persistently activate the RAS signaling pathway, promoting tumorigenesis, while also engaging the PI3K pathway, which foster cell survival, growth, and therapy resistance [63]. Recent studies also implicate the GTF3C6 and FAK pathway in the progression of KRAS-driven LUAD, associating them with enhanced cell migration and invasion. Additionally, KRAS mutations alter cholesterol efflux, which may further contribute to tumor growth [63]. These mutations also influence the tumor microenvironment and immune responses, potentially impacting the success of immunotherapies [64].

The prevalence of MET overexpression in NSCLCs varies significantly, with reported rates between 15% and 70%. Both MET mutations and amplifications act as key oncogenic drivers and therapeutic biomarkers in LUAD [65]. The MET signaling pathway is activated when hepatocyte growth factor (HGF) binds to the MET receptor, initiating downstream signaling cascades that promote cell proliferation, survival, and motility, all of which are crucial for tumor growth and metastasis [66, 67]. In Fak, MET activation enhances cell growth, survival, and movement by stimulating the RAS, RAC, PI3K, ERK, and CAS-CRK pathways. Additionally, MET signaling facilitates invasion, enabling cells to break down or reorganize the surrounding matrix and move across tissue barriers, often utilizing urokinase-type plasminogen activator (uPA), plasminogen activator inhibitor-1 (PAI-1), and matrix metalloproteases (MMPs) [66]. Moreover, alterations in MET are linked to resistance against EGFR tyrosine kinase inhibitors, highlighting the necessity for combination therapies that target both pathways [68].

While the exact prevalence of aberrant mTOR expression in lung cancers is unclear, phosphorylated mTOR has been detected in over 70% of 110 tested NSCLC samples [69]. The mTOR pathway plays a critical role in LUAD, particularly through the PI3K/AKT/mTOR pathway, which stimulates cell proliferation, survival, and metabolism [70]. This pathway is often activated by mutations in upstream regulators such as KRAS. Additionally, mTOR regulates autophagy in LUAD, suppressing it to promote tumor growth, while inhibiting mTOR can enhance autophagy and improve responses to chemotherapy [71]. mTOR signaling also affects the immune response within the tumor microenvironment, influencing tumor progression and immunotherapy outcomes [70].

The TP53 gene serves as a crucial tumor suppressor that regulates various cellular pathways significantly impacted in LUAD, with mutations found in about 25-50% of NSCLC patients [72]. These mutations interfere with cell cycle regulation, especially at the G1/S checkpoint, resulting in unchecked cell proliferation and tumor progression [73, 74]. Additionally, TP53 is critical for triggering apoptosis in response to DNA damage. Mutations in this gene hinder this process, leading to resistance against therapies [74, 75]. TP53 is also involved in the DNA damage response and is associated with the activation of the PI3K/AKT/mTOR pathway, which supports cell survival and growth [73]. Moreover, TP53 mutations can modify the tumor microenvironment and influence immune responses, potentially improving the efficacy of immunotherapies, while also disrupting other signaling pathways such as E2F and Notch, which further exacerbate the aggressive nature of LUAD [73, 76].

FGFR2 mutations occur in about 4-5% of patients with NSCLC, including those with LUAD [77]. The fibroblast growth factor receptor 2 (FGFR2) signaling pathway plays a vital role in the development and progression of LUAD by facilitating cell proliferation and migration, especially in high-grade tumors [78]. Activation of this pathway triggers the PI3K/AKT and MAPK cascades, enhancing cell survival, proliferation, and invasion. It also engages the c-Jun-YAP1 axis, regulating key genes involved in cell growth and survival [78]. Additionally, FGFR2 alterations frequently coincide with changes in the PI3K pathway in LUAD, suggesting that combination therapies targeting both pathways could be an effective strategy for treating FGFR2-positive LUAD [61].

Additional biomarkers identified for LUAD include mutations in PIK3CA [79], BRAF, EGFR, ERBB2[80] and FGFR3 [81], MTOR, and TP53, highlighting the diverse molecular landscape of the disease.

After analyzing the gene-miRNA regulatory network, miR-335-5p, miR-16-5p, miR-125b-5p, miR-21-5p, miR-17-5p, miR-27a-3p, miR-124-3p, miR-26b-5p, miR-155-5p, miR-125a-5p emerged as the top 10 miRNAs based on their degree of connectivity within the network.

miR-335-5p exhibits dual roles, acting as both an oncogenic and tumor suppressor, depending on its target. In several cancers, like prostate cancer, it suppresses cell proliferation, whereas in LUAD, it can promote metastasis. one of its oncogenic roles is enhancing migration by targeting copine-1 (CPNE1), an NF-κB signaling suppressor, through binding to the 3′UTR [26]. Additionally, miR-335-5p downregulates CCNB2, a G2/M phase cyclin that is highly expressed in LUAD by targeting its 3’-UTR [27].

miR-16-5p plays a complicated role in LUAD, with both upregulation and downregulation reported in studies. While circulating miR-16-5p can be upregulated in LUAD and show high sensitivity and specificity for lung cancer prediction in plasma [29], it also has been mentioned to function as tumor suppressor in radioresistant lung cancer cells by regulating key signaling pathways and associating with apoptosis [82]. Exosomal miR–16-5p overexpression reduces PD- L1 expression, which otherwise contributes to tumor growth [30]. Future studies are needed to clarify these contradictory roles in LUAD.

miR-125b-5p is downregulated in LUAD, contributing to tumor proliferation and migration. It has been shown to inhibit cell proliferation and induce apoptosis, emphasizing its tumor-suppressive function [31].

miR-21-5p is upregulated in LUAD, promoting metastasis. It negatively regulates genes like CREBRF, HGF, LIFR, PIK3R1 and XKR6, with PIK3R1being its primary target [33]. It also inhibits the PTEN gene and reduces the expression of apoptosis-related genes such as caspase-9, BAD, and P53 [34].

miR-17-5p is overexpressed in LUAD and is involved in tumor growth and metastasis. It negatively co-expresses with PLSCR4 (phospholipid scramblase 4), a protein involved in immuno-activation, tumorigenesis, apoptosis [35]. Moreover, miR-17-5p has negative correlation with HCP5 and HOXA7 [83]. While primarily oncogenic, some studies suggest that miR-17-5p may act as a tumor suppressive role in early-stage cancers. Further research is necessary to understand its context-dependent functions [36].

miR-27a-3p is elevated in NSCLC tissues but reduced in pulmonary macrophages and peripheral blood [84]. Although miR-27a-3p has been identified in multiple cancer types, the precise mechanisms and signaling pathways in which it influences tumor development, invasion, and metastasis are still not well understood [37]. Recent studies highlight miR-27a-3p has significant role in processes such as epithelial-mesenchymal transition(EMT), immune responses, and chemotherapy resistance. It modulates critical pathways, including Ras/MAPK/ERK pathway, AKT pathway, and TGF-β pathway, affecting oncogenic proteins like c-myc, PI3K, and SMAD2/4. Furthermore, miR-27a-3p activates the Wnt/β-catenin pathway, further promoting cancer cell proliferation and survival [37].

miR-124-3p is significantly downregulated in NSCLC compared to healthy individuals [38]. This miRNA inhibits metastasis by blocking the PI3K/AKT pathway and targeting the disintegrin and metalloproteinase 15 (ADAM15), preventing cancer cell invasion [39]. It also regulates cell migration by through ZEB1 suppression and regulates proliferation by targeting oncogenic pathways involving CD164 and Cadherin-2 (CDH2) [38]. Low levels of miR-124-3p have been associated with gefitinib resistance in NSCLC patients, while increasing its expression can reverse this resistance by inhibiting SNAI2 and STAT3 [38]. Moreover, miR-124-3p influences cellular metabolism, autophagy, and cell cycle regulation, further emphasizing its function as a tumor suppressor in LUAD [38].

miR-26b-5p is downregulated in LUAD tissues, and its increased expression is associated with enhanced radiosensitivity and apoptosis in LUAD cell lines like A549 cells [41]. It has been documented that silencing DUXAP8 expression leads to decreased cell proliferation and increased apoptosis in LUAD by targeting miR-26b-5p, which functions as a cancer promoter [40]. Furthermore, ATF2 has been introduced as a target of miR-26b-5p; when ATF2 is overexpressed, it reduces radiosensitivity, indicates it as a potential biomarker for discriminating cancer cell from normal cells [41]. This miRNA is crucial in various biological processes, such as hypoxia and inflammation, and it enhances cancer cell survival by inhibiting the von Hippel– Lindau (VHL) tumor suppressor protein [42].

miR-155-5p aids in cancer progression by facilitating EMT and plays a role in the regulation of glucose metabolism and immune responses [42]. It is a key regulator of multiple cancers pathways that contribute to tumor growth and survival [42].

miR-125a-5p functions as a tumor suppressor in LUAD cells leads to a decrease in cell viability, proliferation, and invasion, while enhancing apoptosis [44, 45]. It was reported that miR-125a- 5p binds to the 3′-UTR of STAT3, resulting in lower STAT3 expression, which is associated with cancer development [44]. Furthermore, miR-125a-5p specifically targets NEDD9, a protein linked to invasive characteristics, and its overexpression diminishes the invasive potential of the A549 and H1299 lung cancer cell lines [45].

Among these miRNAs, all except miR-17-5p, miR-27-5p have been identified as LUAD biomarkers. While both miR-17-5p and miR-27a-3p play significant roles in LUAD pathogenesis, the term “biomarker” has not been explicitly applied to them in the context of LUAD. This distinction suggests that further research may be required to clarify their potential use as diagnostic or prognostic markers in LUAD.

## Conclusion

In this study, we obtained 48 genes from eight different databases and further refined them to highlight key hub genes, top miRNAs and most significant molecular mechanisms involved in LUAD pathogenesis. The analysis revealed that several of these genes exhibited mutations compared to normal lung tissue, with corresponding miRNAs displaying either upregulation or downregulation in specific cases. We propose that these hub genes and miRNAs are crucial in the initiation and progression of LUAD tumors.

Through a novel approach combining network biology methods and database mining, we found core genes, pathways, and miRNAs linked to LUAD. While several miRNAs were already known as biomarkers, we suggest miR-17-5p and miR-27a-3p as novel biomarkers for LUAD, meriting further experimental investigations.

Overall, our findings offer a concise but impactful subset of molecular drivers in LUAD. The gene prioritizations techniques used in this study streamline the selection of important genes and miRNAs, providing a focused set for future in vivo and in vitro studies, ultimately saving time and resources in experimental validation.

## Data Availability

All data produced are available online at dbGaP, TCGA, Disgenet, OpenTarget, UniProt, CancerHotspot, and ClViC databases.

## Notes

### Competing Interest Statement

The authors have declared no competing interest.

### Funding Statement

This study did not receive any funding

### Author Declarations

The study used (or will use) ONLY openly available human data that were originally located at: dbGaP, TCGA, Disgenet, OpenTarget, UniProt, CancerHotspot, and ClViC databases.

